# TXNIP is a critical regulator of human cardiometabolic health

**DOI:** 10.1101/2025.06.09.25328488

**Authors:** Julia-Josefine Scholz, Sharlaine Y.L. Piel, Ioannis Evangelakos, Christian Müller, Daniela Karall, Birgit Reinhart-Steininger, Katja Steinbrücker, Thomas Stulnig, Julia Rohde, Johannes Mayr, Matias Wagner, Ali Biabani, Thomas Mair, Kilian Müller, Hartmut Schlüter, Sören Weidemann, Viktoria Chirico, Christina Weiler-Normann, Christian Hagel, A.S. Knisely, Joerg Heeren, Benedikt Schoser, Christian Kubisch, Maja Hempel, Holger Prokisch, Saskia B. Wortmann, Anna Worthmann, Christian Schlein

**Author notes:** These authors contributed equally to this work (Julia-Josefine Scholz, Sharlaine Y.L. Piel).

## Abstract

Thioredoxin interacting protein, encoded by *TXNIP*, is a key mediator of glucose and lipid metabolism in preclinical models. However, its role in human metabolism is not well understood. We have characterized a cohort of individuals with biallelic pathogenic variants in *TXNIP* who exhibit lactic acidosis, dyslipidemia, adult-onset severe cardiomyopathy, hypoglycemia, and skeletal muscle weakness. Mechanistically, TXNIP dysfunction was associated with increased fatty acid synthesis and complex rearrangements of the cellular lipidome and proteome. Restriction of dietary carbohydrates resulted in partial rescue of fatty acid synthesis markers and lipid storage in hearts of *Txnip*-KO mice, but led to dyslipidemia. Our studies show that TXNIP is an important modifier in cardiac and skeletal muscle as well as in lipoprotein metabolism and that biallelic pathogenic variants in *TXNIP* lead to a complex disease affecting cellular lipid metabolism of multiple organ systems with potentially fatal adult-onset cardiomyopathy.

## Introduction

Thioredoxin (TXN) is a central regulator involved in cellular redox homeostasis. During oxidative stress TXN interacts with thioredoxin interacting protein, encoded by *TXNIP*^1^, with translocation of TXNIP to mitochondria, thereby regulating cellular glucose metabolism^2^. Mechanistically, TXNIP supports endocytosis of glucose transporter 4 (GLUT4, encoded by *SLC2A4*) and other glucose transporters^3,4^. This limits glucose influx in the fasted state. In line, loss of TXNIP thus results in uncontrolled glucose influx and hypoglycemia^5^. In consequence, TXNIP influences the onset and progression of type 1 diabetes mellitus (DM) and type 2 DM (T2DM)^6, 7^and its pharmacological inhibition has been investigated as a promising target in combating T2DM. However, preclinical studies showed that TXNIP deficiency in mice results in dyslipidemia^8^. This has prompted questions about the role of TXNIP in cellular and systemic lipid metabolism.

The mouse line HcB-19/Dem (HcB-19) is associated with hypertriglyceridemia and hypercholesterolemia. Interestingly, it carries a spontaneous mutation in *Txnip*^8^ (initially named *Hyplip1*^9^). In *Txnip*-knockout (KO) mice, levels of SREBP-1c, a transcription factor mediating the expression of genes associated with lipid synthesis, are higher than wild type littermates, implicating Txnip deficiency in higher rates of hepatic fatty acid synthesis^9^. As fatty acid synthesis is important for the development of dyslipidemia and hepatic steatosis^10^, to speculate that *de novo* fatty acid synthesis reflects the mechanistic connecting link between hypertriglyceridemia and TXNIP^11^ is tempting.

In humans, biallelic pathogenic variants in *TXNIP* have been associated in 3 siblings with mitochondrial disease manifest as lactic acidosis and respiratory chain dysfunction with increased ketone body synthesis. One child had hypoglycemia and elevated serum transaminase activity, hinting at hepatic involvement. However, cardiometabolic (pre-) clinical or mechanistic investigations were not performed^13^.

Using clinical in-depth phenotyping and preclinical models, we have expanded the spectrum of *TXNIP*-associated disease, highlighting its association with an adult-onset cardiomyopathy and hypertriglyceridemia, with variable manifestations of developmental delay and seizures. Mechanistically, we found that biallelic variants – nonsense variants, missense variants, or the 1q21.1 thrombocytopenia-absent radius (TAR) deletion – result in high levels of fatty acid synthesis markers in humans, as also seen in *Txnip*-KO mice. Therapeutic investigations in preclinical models revealed elevated fatty acid synthesis and pathological lipid storage in hearts of *Txnip*-KO mice, changes partially reversible by feeding a low-carbohydrate diet. In one patient sugar restriction led to increased self-reported well-being, whereas in another patient a ketogenic diet resulted in severe hypertriglyceridemia. This indicates that dietary sugar and fat intake may importantly modify the clinical phenotype in TXNIP deficiency. In sum, we describe *TXNIP* as a clinically relevant regulator of human and mouse cellular, systemic, and multi-organ lipid metabolism important for cardiovascular health.

## Results

### Clinical characterization of individuals with pathogenic biallelic variants in *TXNIP*

As TXNIP function is associated with glycemic control and lipid homeostasis in preclinical models, we were interested in the role of TXNIP in human lipid metabolism. For this purpose, we identified humans with biallelic variants in *TXNIP* (NM_006472.6), performed in-depth clinical characterization of these individuals (Table 1, individuals #1-6, Fig. 1a; see Supplemental information for medical history), and compared their clinical phenotypes to those of the previously described siblings homozygous for the p.Gln58delinsHisTer variant^12^ (Table 1, Fig. 1a, individuals #7-9). The 6 additional individuals with biallelic variants were from 5 independent families. Three families were recruited within the German / Austrian network for mitochondrial disorders (mitoNET) and 2 families presented individually at medical-genetic policlinics with cardiomyopathy (Table 1). In this cohort, 2 individuals were children (individuals #3 and 6) and 4 were adults (individuals #1, 2, 4, and 5). Patients #3, 4, and 6^13^ had homozygous frameshift variants (Table 1, Fig.1a), #5 had a homozygous missense variant (individual #5), and 2 brothers, #1 and 2, had a TAR deletion in combination with a nonsense variant on the non-deleted allele. Individual #2 died years before medical-genetic evaluation, with testing performed *post mortem*.

**Fig. 1:**
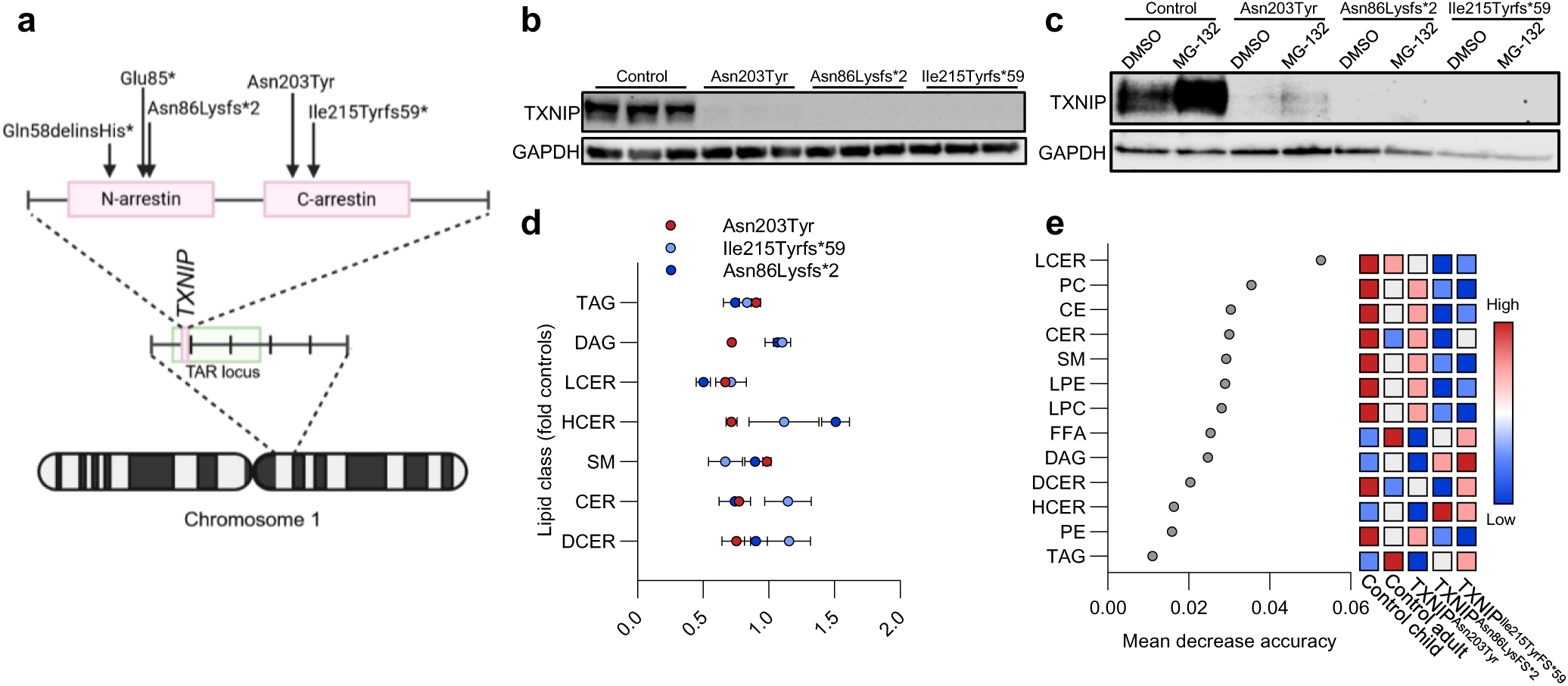
Biallelic variants in *TXNIP* eliminate TXNIP from patient-derived fibroblasts. (a) Chromosomal location and structure of *TXNIP* with C- and N-terminal arrestin domains. Arrows indicate variant position and genes included in the TAR deletion. (b) Western blot, lysates, control fibroblasts (n=3) and TXNIP Asn203Tyr, Asn86Lysfs*2, and Ile215Tyrfs*59 fibroblasts (each n=3), against TXNIP and GAPDH (loading control). (c) Western blot, lysates as above, against TXNIP and GAPDH after treatment with 10 mM MG-132. (d) Relative distribution of lipid classes in TXNIP Asn203Tyr, Asn86Lysfs*2, and Ile215Tyrfs*59 fibroblasts. (e) Random forest plot of lipidome, TXNIP Asn203Tyr, Asn86Lysfs*2, and Ile215Tyrfs*59 fibroblasts, compared to healthy controls (n=5). TAG = triacylglycerides, DAG = diacylglycerides, LCER = lactosylceramides, HCER = hexosylceramides, SM = sphingomyelins, CER = ceramides, DCER = dihydroceramides.

**Table 1:**
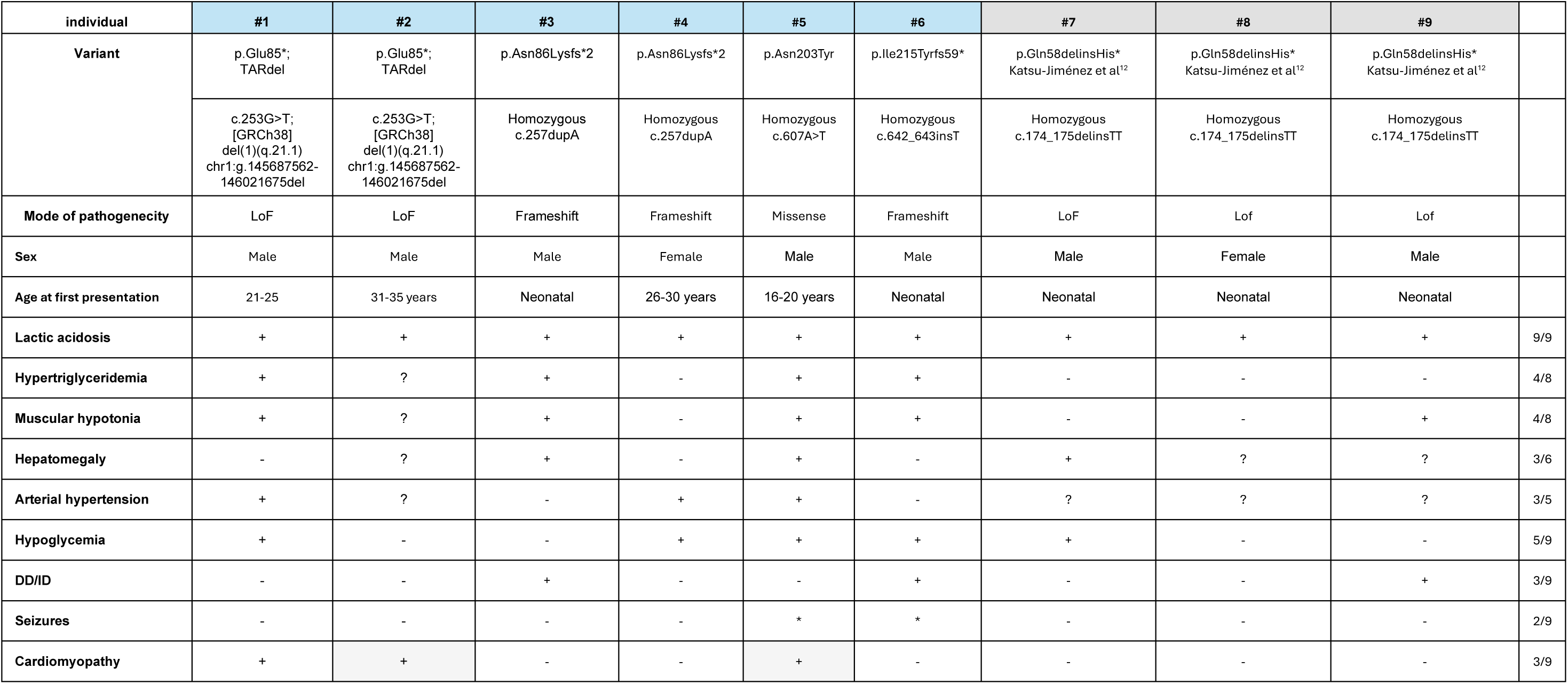
Clinical table of individuals with biallelic variants in *TXNIP*. Nine patients, 6 novel (from 5 families) and 3 reported, are included. The most frequent features of disease were lactic acidosis (9/9), hypoglycemia (5/9), hypertriglyceridemia (4/8), and muscular hypotonia (4/8). Cardiomyopathy (adult-onset) was present in 3 patients, leading to death in 2 (grey background). Patients #1 and #2 were brothers. LoF, loss of function; DD/ID, developmental delay / intellectual disability

The main clinical feature (cf. Supplemental information) in the 3 individuals described by Katsu-Jiménez et al.^12^ and the 6 newly studied individuals was lactic acidosis (present in 9/9). Hypoglycaemia (5/8 evaluated individuals), hypertriglyceridemia (4/8), muscular hypotonia (4/8), arterial hypertension (3/5), hepatomegaly (3/6), cardiomyopathy (3/9), developmental delay (3/9), and seizures (2/9) also were observed. Lactic acidosis was present on every routine clinical-chemistry assessment; although hypertriglyceridemia was repeatedly seen, triglyceride values were occasionally within the reference range in some individuals. Three of 4 adults had severe complex cardiomyopathy. Individual #2 died in his 30’s of cardiomyopathy-associated complications one week after cardiomyopathy was diagnosed. Individual #5 died in his 30’s after an anaesthesia-associated cardiac arrest. He had severe hypertriglyceridemia (plasma triglycerides > 1600 mg/dl, ref. < 150 mg/dl). Individual #6, normolipidemic at birth, developed hypertriglyceridemia (≤ 364 mg/dl). Individual #3 had mild hypertriglyceridemia, which was exacerbated on a ketogenic diet (> 10,000 mg/dl). Impaired function of TXNIP thus appears to affect especially muscle- and liver-related functions, deranging metabolic homeostasis and leading to dyslipidemia, hypoglycemia, and lactic acidosis.

### Biallelic variants in *TXNIP* eliminate TXNIP from patient-derived fibroblasts

To assess the impact of different *TXNIP* variants on TXNIP abundance (Fig.1a), we performed western blot analysis of cell lysates of cultured patient-derived fibroblasts. The results confirmed that TXNIP was not detectable in lysates of fibroblasts derived from individuals with a p.Asn86Lysfs*2 or a p.Ile215Tyrfs*59 variant in *TXNIP* (Fig.1b). However, in the fibroblasts derived from individual #5, with a p.Asn203Tyr variant in *TXNIP*, a residual minimal fraction of TXNIP was detected (Fig.1b). We hypothesized that the protein is translated but rapidly degraded via the ubiquitin-proteasome pathway as a result of protein quality control. To investigate this, we incubated all patient-derived fibroblasts with the proteasome inhibitor MG-132 and performed western blot analysis. A signal was detected in fibroblasts from individual #5, indicating that the encoded protein undergoes rapid posttranslational degradation (Fig.2c). Lipidome analyses (“lipidomics”) from patient-derived fibroblasts from individuals #4-6 showed minor alterations, mostly distinguished from controls by low lactosyl-ceramide levels (LCER) in random forest plot analysis (Metaboanalyst^14^). Low levels of LCER, a lipid class involved in redox stress^15^ (Fig.1d+e), might indicate that *Txnip* deficiency reduces redox stress^16,17^ in this metabolically rather inactive setting. However, as the clinical phenotype mostly involved metabolically active tissues and the circulation, we further focussed on metabolically active organs and related models for further clinical and preclinical investigations.

**Fig. 2:**
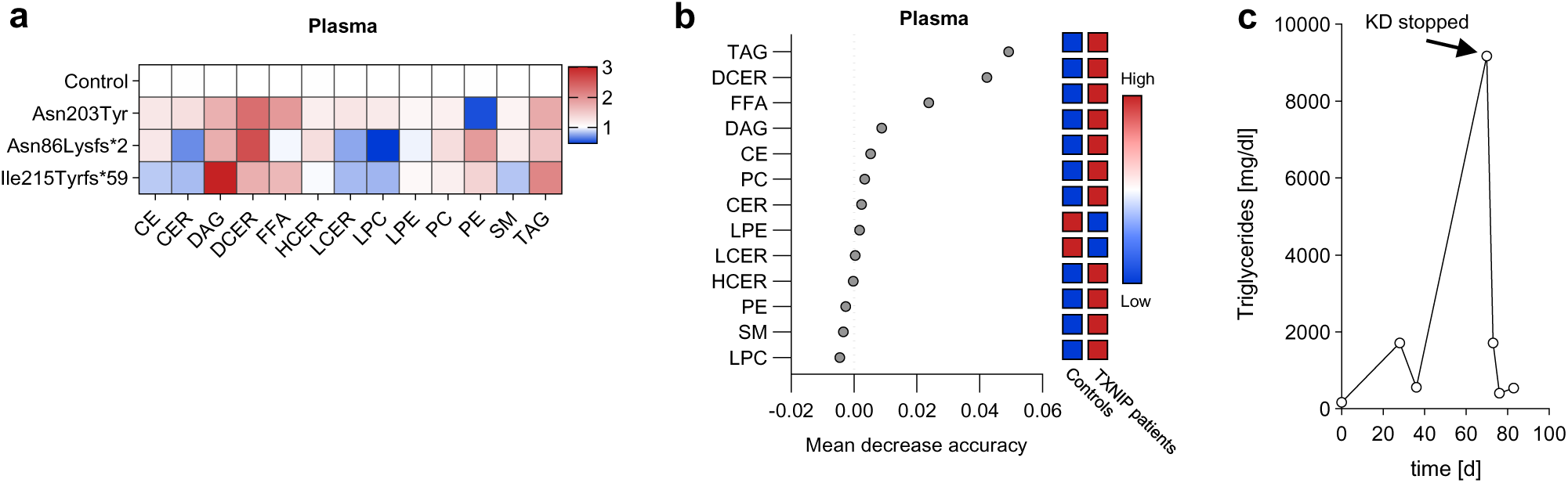
Complex lipidome rearrangements in plasma of individuals with biallelic variants in *TXNIP.* (a) Heatmap of plasma lipid concentration -fold changes, patients with TXNIP variants Asn203Tyr, Asn86Lysfs*2, and Ile215Tyrfs*59 (each n=1) versus controls (n=9). (b) Random forest plot of lipid class concentration -folds, plasma of patients (n=3) versus controls (n=12), as mean decrease accuracy. (c) Plasma triglyceride measurements before, during, and after start and stop of ketogenic diet, individual #3.

### Complex lipidomic rearrangements in plasma of individuals with biallelic variants in *TXNIP*

The association of *TXNIP* dysfunction with hypertriglyceridemia, described in *Txnip*-KO mouse models, was also seen in a subset of individuals with biallelic variants in *TXNIP.* To further characterize the impact of *TXNIP* dysfunction on human lipid metabolism, we performed targeted lipidome analysis of the plasma of patients and of age- and sex-matched healthy controls. We detected over 800 lipid species from 13 lipid classes. Random forest plotting of lipid class concentrations (Metaboanalyst^14^) revealed that with biallelic pathogenic variants in *TXNIP* triacylglycerides (TAG), diglycerides (DAG), and free fatty acids (FFA) were the top lipid classes, distinguishing individuals with such variants from their matched controls (Fig.2a). Interestingly, biallelic variants in *TXNIP* were also associated with higher levels of dihydroceramides (DCER), a lipid class important in cellular stress responses: Plasma levels of DCER have been connected to metabolic disease^18^ (Fig.2a,b). Elevations in fasting TAG were mild to moderate (around twice those in controls), but were detected in all 3 available patient-derived plasma samples. Additionally, levels of DAG in patients were higher than in controls, suggesting either hydrolysis of TAG or a decreased fatty acid re-esterification rate within the liver and subsequent release of immature complex lipids into plasma lipoproteins.

Speciation of DAG showed higher abundance of saturated lipids, such as those with 2 palmitate residues, DAG(16:0/16:0); with palmitate and oleate, DAG(16:0/18:0); or the rare di-arachidin, DAG(20:0/20:0) (Fig.S1a). Overall, in light of the higher saturated fatty acid residues, the question arose whether endogenous fatty acid synthesis, a major source of saturated fatty acids^19^, led to elevated TAG or if elevated TAG resulted from higher cellular lipid uptake.

Of note: Consuming a ketogenic diet, individual #3 developed severe hypertriglyceridemia (peak >10,000 mg/dl, Fig.2c), which rapidly reversed after that diet ended. This indicates that diet is a major regulator of hypertriglyceridemia in individuals with biallelic variants in *TXNIP*.

### TXNIP as a regulator of metabolic pathways in muscle

Transmission electron microscopy (TEM) of striated muscle of individual #5 revealed accumulation of lipid droplets and glycogen deposits (Fig.3a) more abundant than in controls. We assessed the composition of the lipid deposits by targeted lipidome analyses. In line with the fact that TAG are the predominant form of stored lipids in cellular lipid droplets, we found high levels of TAG in the muscle of individual #5. Further, levels of hexosylceramides (HCER), DCER, and ceramides (CER) were slightly elevated, whereas levels of cholesteryl esters, phosphatidylcholine, phosphatidylethanolamine, and sphingomyelin were lower (Fig.3b). These findings indicated robust and complex remodelling of muscle. When we analysed the concentration of specific lipid species, TAG containing saturated or monounsaturated fatty acids in particular were mostly elevated (Fig.3c). The absolute concentration of saturated, monounsaturated, and polyunsaturated fatty acids was elevated, with saturated and monounsaturated fatty acids as the main fraction.

**Fig. 3:**
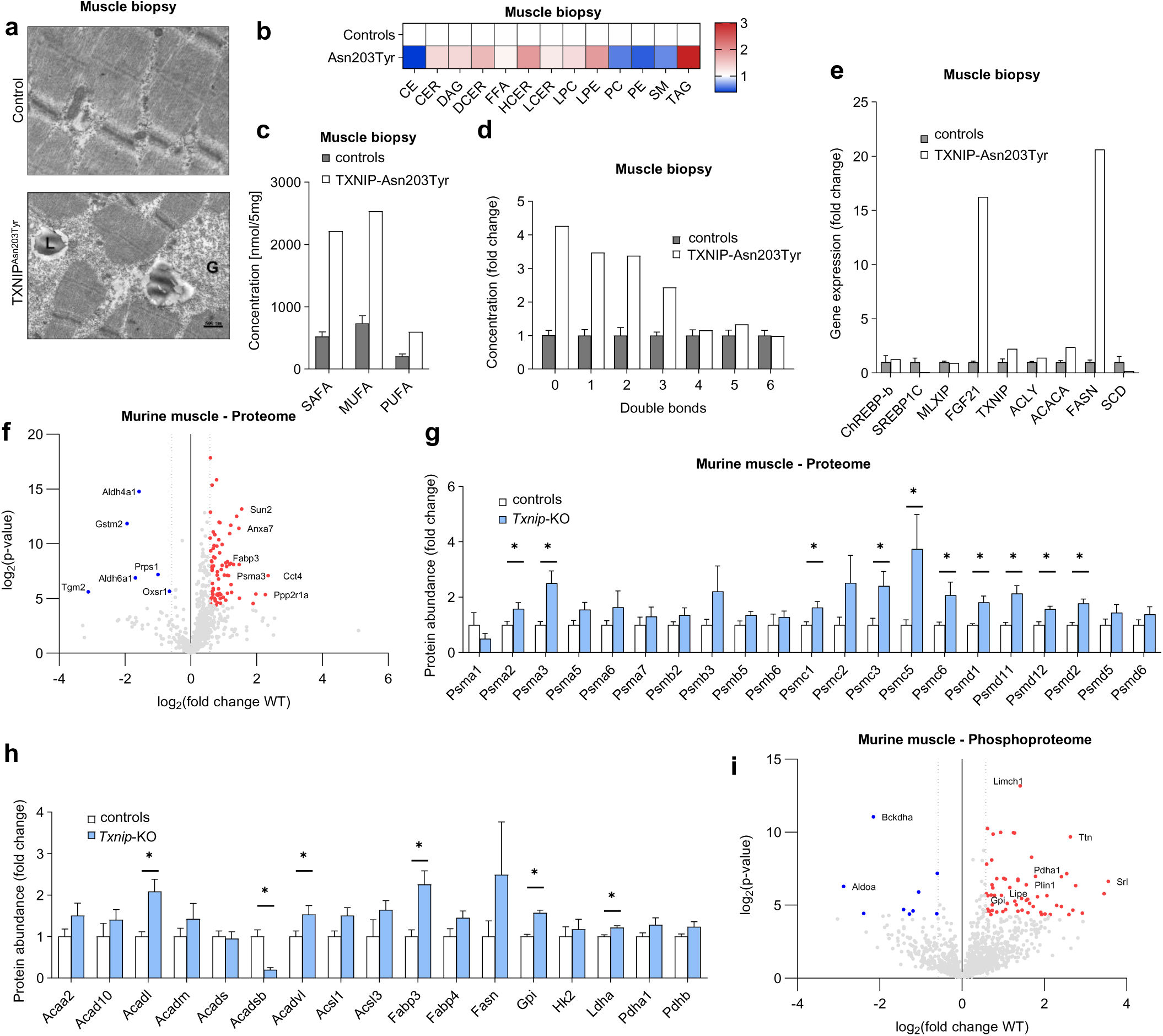
TXNIP as a regulator of metabolic pathways in skeletal muscle. (a) TEM images, control muscle tissue and muscle biopsy specimen, individual #5. (G), glycogen; (L), lipid droplets. (b) Heatmap showing muscle lipid class concentration -fold changes, muscle biopsy, individual #5 (n=1) versus controls (n=3). (c) Concentrations of saturated fatty acids (SAFA), monounsaturated fatty acids (MUFA), and polyunsaturated fatty acids (PUFA), muscle biopsy specimen, individual #5 (n=1) versus controls (n=3). (d) Fold change, concentration of double bonds in fatty acids, muscle biopsy specimen, individual #5 (n=1) versus controls (n=3). (e) Gene expression-fold changes, key regulators in muscle biopsy specimen, individual #5 (n=1) versus healthy controls (n=3). (f) Volcano plot and (g-h) protein abundance (proteomics), muscle of *Txnip*-KO mice and of WT littermates. (i) Volcano plot, phosphoproteomics, muscle of *Txnip*-KO mice and of WT littermates.

Plotting of double bonds of fatty acid residues within the TAG showed that lipid accumulation comprised TAG species with low levels of desaturation or without desaturation (Fig.3d). This composition exists in tissues with very high fatty acid synthase activity, which prevents the use of polyunsaturated fatty acids for complex lipids^10^. Gene-expression analysis in muscle of individual #5 revealed higher expression of *ACACA* (2.3-fold) and *FASN* (20-fold), key enzymes in the *de novo* fatty acid synthesis pathway, as well as stress-related *FGF21* (16-fold) (Fig.3e). However, expression levels of canonical transcription factors of endogenous fatty acid synthesis in skeletal muscle, such as *MLXIP* or *SREBP1c*, were not altered. Moreover, expression of the desaturase stearoyl-CoA 9-desaturase (*SCD*) was lower. SCD catalyses mono-unsaturated fatty acid production and is usually transcriptionally co-expressed with *FASN*, indicating that the increase in MUFA might be also partially driven by uptake of exogenous lipids from the circulation.

To generalize these findings obtained in a biopsy specimen from a single patient, we performed untargeted proteome analyses (“proteomics”) of skeletal muscle from wild-type (WT) and *Txnip*-KO mice (Fig.3f-h). In *Txnip*-KO murine muscle, proteasome subunits and regulators of the PSMA/PSMB/PSMC/PSMD families were substantially upregulated, possibly indicating derangement of the proteasome (Fig.3g). Lipid metabolism, which was widely affected in humans, showed a trend towards higher pathway translation, such as higher ACADL, ACADVL, and FABP3 and a trend to higher FASN in *Txnip*-KO mice (Fig.3h). Phosphoproteome analyses (“phosphoproteomics”) of skeletal muscle of WT and *Txnip*-KO mice, conducted to investigate underlying signaling pathways, indicated elevated levels of lipid-droplet – associated proteins (PLIN1, LIPE), and reduced 2-oxoisovalerate dehydrogenase subunit alpha (BCKDHA), active in branched-chain amino acid metabolism, and pyruvate dehydrogenase (PDHA1) (Fig.3i). Thus, TXNIP thus proved to affect multiple anabolic and catabolic pathways in skeletal muscle, such as proteostasis and lipid metabolism.

### Elevated liver fatty acid synthesis in an individual with biallelic variants in *TXNIP*

As hepatomegaly was noted in some individuals with biallelic variants in *TXNIP*, we investigated the effects of TXNIP deficiency on the liver. Individual #5 underwent liver biopsy because of elevated serum transaminase activity. Both light microscopy (haematoxylin / eosin, H&E; periodic acid – Schiff technique, PAS) and TEM of the liver-biopsy specimen revealed a moderate accumulation of glycogen together with lipid droplets (Fig.4a), as observed for muscle (Fig.3a). Tissue had been asserved; gene-expression analysis found higher expression of genes related to *de novo* fatty acid synthesis, including *FASN* and *SCD* (both 5-fold), as well as of key lipogenic transcription factors such as *ChREBP-beta* and *SREBP1c* (Fig.4b). Transcriptional activation of lipogenesis pathways was higher than in obese patient controls (Fig.4b), suggesting a condition already connected with high lipogenesis^11^. Patient material did not suffice for further lipidome analyses. Accordingly, and in hopes of further generalizing these findings, livers from WT and *Txnip*-KO mice were studied. Lipidomics of livers revealed significantly higher levels of CER (Fig.4c) in *Txnip*-KO mice than in WT littermates, a lipid class affected in inflammation^20^ and in insulin resistance^21^. Further, compositional analysis of TAG revealed a significantly higher proportion of saturated fatty acids, potentially indicating increased endogenous fatty acid synthesis activity in *Txnip*-KO livers compared to livers of WT mice (Fig.4d). To gather mechanistic insights into the lipidomic fingerprint of the *Txnip*-KO livers, we performed untargeted proteomic analysis (Fig.4d). A volcano plot analysis revealed low levels of enzymes involved in steroid metabolism (HSD3B3, CYP17A1) but no clear pattern resembling the gene-expression phenotype seen in individual #5. Despite compositional changes supporting high endogenous fatty acid and CER synthesis (Fig.4c), hepatic protein expression in the fatty acid synthesis pathway (ACACA, ACSS2, SCD1) and in CER synthesis (CERS2) was reduced or unchanged in *Txnip*-KO mice, indicating regulation independent of hepatic protein expression.

**Fig. 4:**
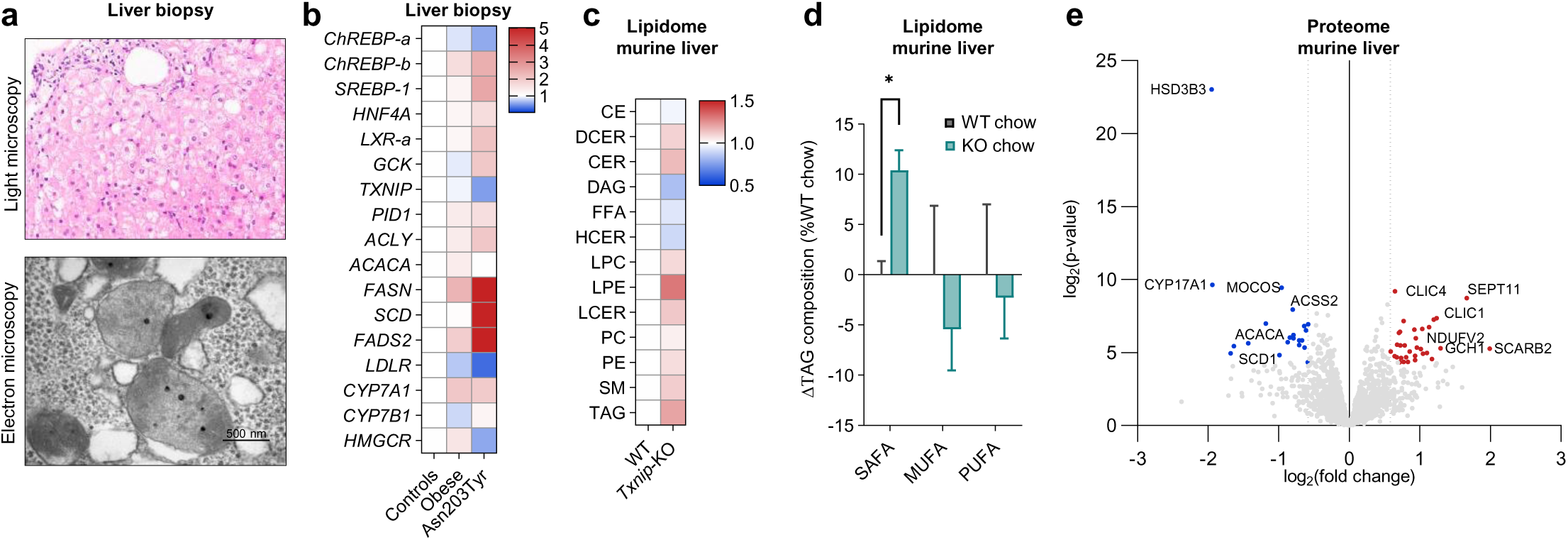
Elevated liver fatty acid synthesis in an individual with biallelic variants in *TXNIP*. (a) Light micrograph (H&E) and (b) TEM image of liver tissue, individual #5, showing glycogen accumulation and abnormal mitochondrial cristae. (b) Heatmap showing gene expression, liver of TXNIP Asn203Tyr patient (n=1) versus healthy controls (n=8) and obese individuals (n=9), red indicating higher and blue lower gene expression. (c) Random forest plot, liver lipidome, *Txnip*-KO mice and WT littermates. (d) Volcano plot, liver proteome, *Txnip*-KO mice and WT littermates. CE = cholesterol esters, CER = ceramides, DAG = diacylglycerides, DCER = dihydroceramides, FFA = free fatty acids, HCER = hexosylceramides, LCER = lysosylceramides, LPC = lysophosphatidylcholine, LPE = lysophosphatidylethanolamine, PC = phosphatidylcholine, PE = phosphoethanolamine, SM = sphingomyelins, TAG = triacylglycerides.

### TXNIP deficiency leads to storage lipid accumulation in heart tissue

Three of 4 adults with biallelic variants in *TXNIP* had severe cardiomyopathy, which clinically led to the death of 2 (see medical history in supplemental information). Light microscopy of H&E-(Fig.5a) and PAS-(Fig.5b) stained myocardial biopsy specimens from individuals #1 and #2 revealed interstitial fibrosis in #1 and myocyte hypertrophy with severe lipomatosis in #2; biopsy was conducted in #2 several days before death.

**Fig. 5:**
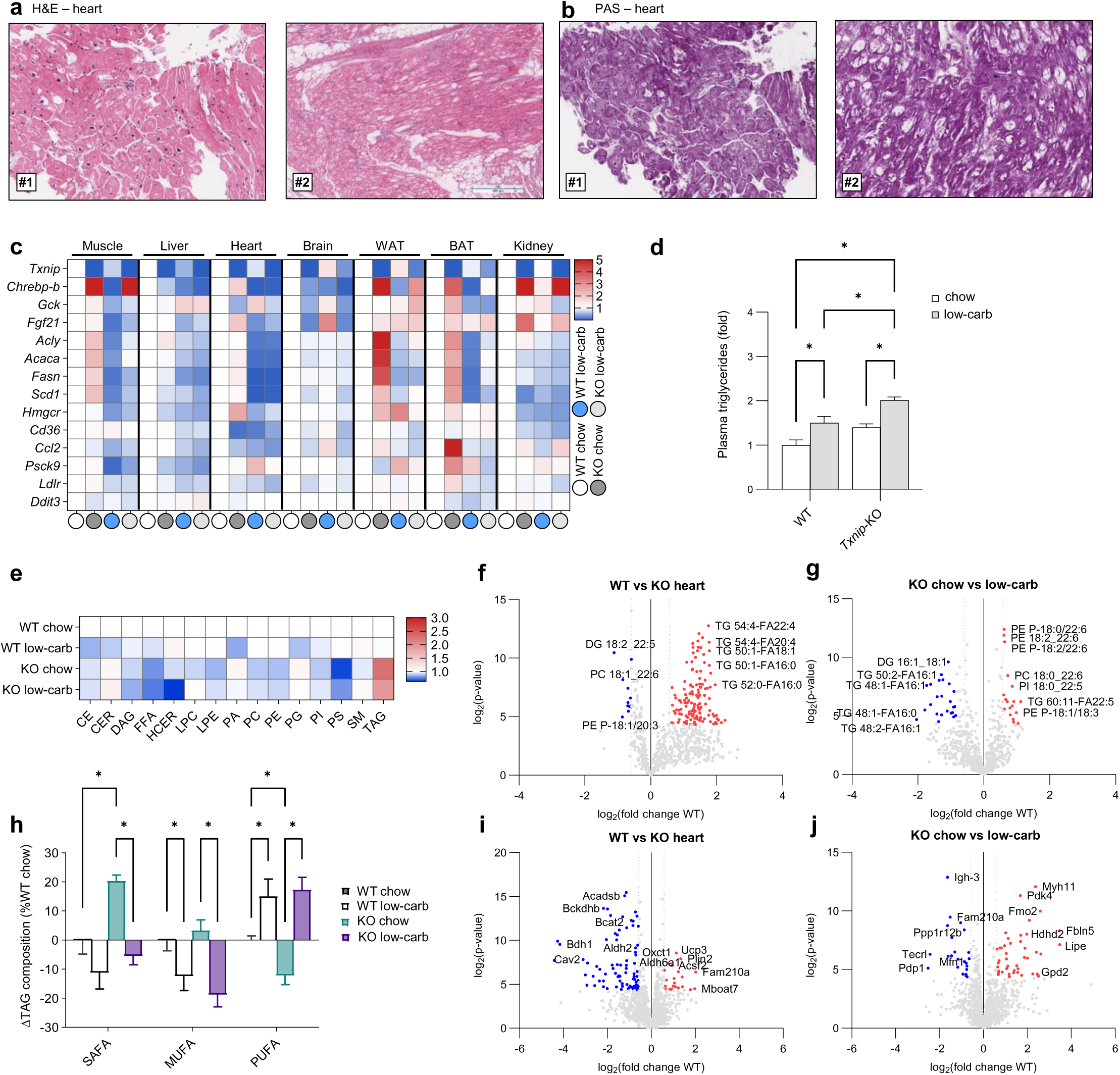
Therapeutic limitation of dietary carbohydrates to alleviate increased lipogenesis. (a), (b) Light micrographs, myocardial biopsy specimens, individuals #1 and #2 ; H&E, (a), and PAS, (b). (c) Heatmaps, gene expression, muscle, liver, heart, brain, white adipose tissue (WAT), brown adipose tissue (BAT), and kidney, *Txnip*-KO and WT mice on standard chow or low-carbohydrate diet (n=5). (d) Plasma triglycerides, *Txnip*-KO and WT mice on chow or low-carbohydrate diet (n=5). (e) Heat map and (f,g) volcano plots and (h) TAG lipid class composition, heart tissue lipidomes, *Txnip*-KO and WT mice on standard chow or low-carbohydrate diet (n=5). (i,j) Heart tissue proteomes, *Txnip*-KO and WT mice on standard chow or low-carbohydrate diet (n=5). CE = cholesterol esters, CER 18:1 = ceramides, DAG = diacylglycerides, DCER = dihydroceramides, FFA = free fatty acids, HCER = hexosylceramides, LPC = lysophosphatidylcholine, PA = phosphatidic acid, PG = phosphatidyl glycerol, PI = phosphatidyl inositol, PS = phosphatidyl serine, LPE = lysophosphatidylethanolamine, PC = phosphatidylcholine, PE = phosphoethanolamine, SM = sphingomyelins, TAG = triacylglycerides.

Interestingly, a detailed anamnesis found that for many years individual #1 had strictly limited sugar intake for several weeks, as to do so improved his own sense of well-being and muscular fitness (see supplemental information). We hypothesized that this phenomenon might be explained by reductions in cellular fatty acid synthesis and sought to mimic this in our preclinical model, studying WT and *Txnip*-KO mice on standard-chow and low-carbohydrate diets.

Gene-expression analysis was used to screen a set of mouse organs for markers of increased fatty acid synthesis. This revealed that *Txnip*-KO mice on a standard-chow diet exhibited higher expression of markers associated with fatty acid synthesis, such as the lipogenic transcription factor carbohydrate responsive element binding protein, isoform beta (ChREBP-beta)^22^, an alternative splicing product encoded by *Mlxipl* and a well-described interaction partner of TXNIP^23^, in skeletal muscle, heart, white adipose tissue, brown adipose tissue, and kidney (Fig.5c). This was accompanied by higher *Fasn* expression in skeletal muscle, heart, and white and brown adipose tissues (Fig.5a). Additionally, expression of the glycogen synthesis marker *Gck* was notably higher, particularly in the liver, where it represents the predominant isoform, alongside hexokinase 1 and hexokinase 2, which are the main isoforms in other organs (Fig.5c). A low-carbohydrate diet effectively lowered *Fasn* expression in skeletal muscle, heart, and white and brown adipose tissues and lowered Chrebp-beta expression in all organs except kidney and skeletal muscle; in skeletal muscle, instead of ChREBP-beta, *Mlxip,* encoding MondoA, is the main lipogenic transcription factor (Fig.5c).

Overall, based on gene expression of the surrogate marker *Fasn*, a low-carbohydrate diet may be a route to reducing pathologically elevated fatty acid synthesis and, to some extent, glycogen synthesis in *Txnip*-KO mice.

As fatty acid synthesis is hypothesized to be potentially linked directly to the phenotype of elevated plasma TAG levels, we hypothesized that lower fatty acid synthesis might result in lower plasma TAG levels. However, a low-carbohydrate diet resulted in higher plasma TAG levels in both WT and *Txnip*-KO mice (Fig. 5d). This suggests that in *TXNIP* deficiency not high fatty acid synthesis but other mechanisms may drive increases in plasma TAG, such as impaired vascular catabolism of TAG-rich lipoproteins or the fact that to consume a low-carbohydrate diet implicates ingestion of larger amounts of lipids.

As a highly relevant clinical phenotype of *TXNIP* disease was involvement of the heart we also studied the hearts from WT and *Txnip*-KO mice (Fig.5e). *Txnip*-KO mouse hearts displayed elevated expression levels of genes involved in fatty acid synthesis that responded well to a low-carbohydrate diet (Fig.5c). Lipidomics of heart tissues revealed that in *Txnip*-KO mice higher gene expression of the fatty acid synthesis pathway was accompanied by high TAG lipid storage in heart (Fig.5e). It is tempting to speculate that deregulated lipid metabolism also underlies part of the potentially lethal cardiac pathophysiology in humans, as in the myocyte hypertrophy with lipomatosis seen on myocardial biopsy in individual #2 (Fig.5a). That 2 weeks of a low-carbohydrate diet significantly reduced heart TAG storage from values seen with a standard-chow diet (∼20%, *p* < 0.05; Fig.5e) indicates that dietary interventions can lower TAG in heart muscle in *Txnip*-KO mice. Volcano plotting of lipid species found that palmitate species, but also MUFA and PUFA triacylglycerol species, were “top hits” (Fig.5f). However, whilst a low-carbohydrate diet lowered levels of palmitate and palmitoleate TAG species in *Txnip*-KO mice (Fig.5g), it elevated levels of plasmalogens, members of an anti-oxidative lipid class highly abundant in heart tissue^24^, and of polyunsaturated species of phosphatidylethanolamine and phosphatidylcholine (Fig.5f). To compare saturated and unsaturated lipid species quantitatively, TAG composition was plotted for the different groups (Fig.5h). On standard chow, *Txnip*-KO hearts contained more saturated fatty acids and fewer PUFAs than WT hearts. These differences were completely eliminated on a low-carbohydrate diet, indicating that a dietary regimen focused on fatty acid synthesis might be a promising approach to reduce lipid storage and to normalize lipid composition. Proteomic analysis of hearts of *Txnip*-KO mice and WT littermates revealed a complex shift in the proteome of heart tissue in TXNIP deficiency, with increased levels of lipid anabolism marker PLIN2 and reduced levels of markers of branched-chain amino acid metabolism (Fig.5i). Interestingly, a low-carbohydrate diet induced LIPE in *Txnip*-KO mice, indicating that TAG hydrolysis might be a mechanism by which a low-carbohydrate diet reduces lipid storage (Fig.5j).

## Discussion

TXNIP, a multifunctional molecule, is associated with the pathophysiology of diabetes mellitus. It may also serve as a therapeutic target for hyperglycemia and associated diseases. Here, our work shows that biallelic pathogenic variants in *TXNIP*, previously described as causing a mitochondriopathy with hyperlactatemia, can cause adult-onset cardiomyopathy and hypertriglyceridemia that ranges from mild to severe depending on diet.

TXNIP deficiency can result from different types of biallelic nonsense or missense variants in *TXNIP* as well as from a 1q21.1 deletion associated with the TAR syndrome^25^ in combination with variants in *RBM8A*^26^ in *trans*. However, the newly described association of a disease caused by 1q21.1 deletion and compound inheritance with *TXNIP* must be taken into account for all families with TAR deletion.

TXNIP controls cardiac hypertrophy in mice in response to pressure overload^27^. The TXNIP-interaction partner TXN is involved in pressure overload-induced cardiac hypertrophy as well^28^. Individuals with TXNIP deficiency tend to show arterial hypertension (Table 1), suggesting that early control of high blood pressure in TXNIP deficiency may be of special importance. Hearts of *Txnip*-KO mice show increased basal and stimulated insulin signaling^29^ and glucose uptake^30^ as well as inadequate AMP-activated protein kinase fasting responses^31^. These might give rise to higher fatty acid synthesis and lipid storage. Although such lipid storage may be detrimental as part of the pathomechanism causing cardiomyopathy in TXNIP deficiency, it also could underlie the beneficial effect of TXNIP deficiency during myocardial infarction in mice^32,33^, perhaps serving as an increased intracellular energy source during ischemia. Of interest is that TXNIP deficiency protects against high-fat-diet – induced cardiomyopathy^30^, maybe due to the fact that high fat diet reduces endogenous fatty acid synthesis. This might blunt increased lipid storage as a pathogenic effect of TXNIP deficiency, with protective effects outweighing detrimental effects in experimental settings. However, in humans, as constitutive loss of TXNIP seems to lead to a potentially fatal cardiomyopathy, including *TXNIP* variants in (virtual) panel analysis in genetic testing of patients with cardiomyopathy must be recommended.

The association of TXNIP with hypertriglyceridemia in humans has been debated over the years^34–38^. Earlier studies concluded that single-allele variation in *TXNIP* does not cause an autosomal-dominant hypertriglyceridemia. Our work, however, demonstrates that an autosomal-recessive form of hypertriglyceridemia with variable expressivity and incomplete penetrance, most likely depending on additional environmental and dietary factors, results from biallelic variation in *TXNIP*. As we now show the association of hypertriglyceridemia to biallelic variants in TXNIP, routine genetic testing should include variants in *TXNIP* as a rare cause of monogenic autosomal-recessive hypertriglyceridemia.

The role of TXNIP in *de novo* lipogenesis was recognized two decades ago in HcB-19/*Txnip*-KO mice^9^, with higher fatty acid synthesis and re-esterification rates contributing to hepatic steatosis. However, cholesterol synthesis was decreased, whereas the total cholesterol esterification rate was increased. Our work demonstrates that patients with biallelic variants in *TXNIP* can exhibit increased *de novo* lipogenesis-pathway gene expression in liver. Further, gene-expression and lipidome analyses of muscle of a patient with a homozygous putatively pathogenic variant in *TXNIP* revealed highly increased *FASN* expression levels and increased saturated and monounsaturated fatty acids in muscle tissue, possibly leading to lipid deposition and muscle weakness. *De novo* lipogenesis (DNL) in muscle is upregulated in obesity and to cause insulin resistance^39^. Thus, targeting elevated DNL is a promising therapeutic option in patients with biallelic variants in *TXNIP*.

Interestingly, individual #1 reported reduced symptoms during dietary restriction of refined sugar intake, which he imposed on himself for several weeks every year. To determine mechanistically if increased lipogenesis in TXNIP deficiency is derived from dietary carbohydrates, and if this has any impact on the heart, we used a low-carbohydrate diet in *Txnip*-KO mice. Indeed, a low-carbohydrate diet could restore high muscle and heart lipogenesis rates in *Txnip*-KO mice to the lower rates in their WT littermates. Of importance was 1) that low carbohydrate levels in the diet did not exacerbate hypoglycemia in mice (as well as that diet-induced hypoglycemia did not occur in individual #1), and 2) that the diet used was – compared to standard chow – hypercaloric to avoid any calorie restriction effects. However, low carbohydrate intake led to moderately higher plasma triglyceride levels in *Txnip*-KO mice. Further, on a ketogenic diet – low carbohydrate intake with very high lipid intake, frequently used in patients with mitochondriopathies – individual #3 developed severe hypertriglyceridemia. To avoid high pancreatitis risks, dyslipidemia accordingly must be monitored strictly during dietary interventions in *TXNIP* disease.

In sum, we describe TXNIP as a distinct modulator of human lipid and glucose metabolism in a tissue-specific manner affecting the mouse and human lipidome by regulating fatty acid synthesis. Biallelic variants of *TXNIP* are associated with a systemic disease affecting multiple organs and are connected to lactate acidosis, cardiomyopathy, hypoglycemia, hypertriglyceridemia, and muscle weakness.

## Methods

### Ethical statement

All analyses of patient information, plasma samples, and muscle- and liver-biopsy specimens were carried out with written informed consent. Written informed consent for studies in children was obtained from their legally authorized representatives, with consent to publish identifiable information for patients and their parents. All studies were conducted in accordance with the Declaration of Helsinki and approved by the local medical ethics committees (Ethical Review Board of the Ärztekammer Hamburg: PV7038; Salzburg: Mitonet 415-E/1317/13-2020).

### Animal model

All studies on animals were performed with permission of the Animal Welfare Offices of the University Medical Center Hamburg-Eppendorf and the Behörde für Gesundheit und Verbraucherschutz Hamburg. The *Txnip*-KO mouse line used was purchased from Jackson Laboratories. Mice were fed standard chow (16.2 MJ/kg, 67% kJ from carbohydrates) or Ssniff^®^ E15660 (21.2 MJ/kg, 2% kJ from carbohydrates) and had *ad libitum* access to food and water in standard housing conditions. Blood plasma parameters were evaluated using the Cobas c 111 system (Roche). Ritonavir treatment was delivered via intraperitoneal injections. Blood for glucose measurement was taken from the tail, while blood for ethylenediaminetetraacetic acid (EDTA) plasma measurements was taken by cardiac puncture. Collected organs were snap-frozen in liquid nitrogen pending analysis.

### Cell cultures

Fibroblast cultures were derived from patients or from age-matched adult and child controls. Cells were cultured at 37°C in 5% carbon dioxide in Dulbecco’s Modified Eagle Medium with 4.5 g/l D-glucose, L-glutamine, and pyruvate (Gibco, Thermo Fisher Scientific). All media were supplemented with 10% FCS and 1% Anti-Anti (penicillin / streptomycin / amphotericin B; Gibco). To inhibit proteasomal activity, InSolution™ MG132 (Calbiochem, Cat.-No. 474791), 20 µM, was added to cell-culture supernatants for 6 h.

### Gene expression

For RNA isolation, tissues were homogenized in TRIzol^®^ (Invitrogen) using a TissueLyser (Qiagen). The homogenate was mixed with 200 µl of chloroform. The aqueous phase was mixed with 300 µl absolute ethanol and purified using a NucleoSpin RNAII Kit (Macherey-Nagel). RNA concentrations were measured using Thermo Scientific NanoDrop™ equipment. Quality was determined at 260/280 nm absorbance ratio. cDNA was synthesized using a High-Capacity cDNA Archive Kit (Applied Biosystems). Gene expression was measured using SYBR green (Applied Biosystems). The values were normalized to those for the indicated housekeeper.

### Oral glucose tolerance testing

Radiolabeled oral glucose tolerance testing was performed as described^40^. Mice fasted for 4 h were gavaged with glucose (2 g/kg body weight) and of the tracer ^3^H-deoxyglucose (0.72 MBq/kg body weight). Tail-vein blood was collected at indicated time points. Glucose levels were measured using Aviva AccuCheck glucose sticks (Roche). At 2 h, mice were anaesthetized (xylazine/ketamine) and blood for plasma was collected by left ventricular puncture. The mice were then systemic-perfused with PBS-heparin (10 U/ml). Harvested organs were dissolved in 10x (v/w) Solvable (Perkin Elmer) and radioactivity (dpm) was measured by scintillation counting using a Perkin Elmer Tricarb Scintillation Counter.

### Histologic studies

For routine histologic work, harvested organs were fixed in a 4% paraformaldehyde (PFA) solution for at least 24 h. Samples were embedded in paraffin, sectioned, and stained with H&E and PAS techniques. Sections were examined by light microscopy. For ultrastructural study, TEM-fixed tissue was stained *en bloc* with osmium tetroxide and embedded in EPON-812 equivalent Agar 100 Resin (Agar Scientific). Semithin sections counterstained with azure 2 / methylene blue were examined by light microscopy and ultrathin sections stained with uranyl acetate and lead citrate were examined using a Zeiss EM900 transmission electron microscope (Zeiss) and a 2kLJ×LJ2k slow-scan CCD camera (Tröndle Restlichtverstärkersysteme).

For electron microscopic investigation of human muscle biopsy, samples were washed in 0.1 M cacodylate buffer (Sigma-Aldrich), incubated for 2 hours in 1% osmium tetroxide (Science Services, Munich, Germany), dehydrated in an ascending series of ethanol, and embedded in Epon 812 (Serva). Ultrathin sections were counterstained with uranyl acetate (Polyscience, Eppelheim, Germany) and lead citrate (Riedel-de Haën, Seelze, Germany), and analyzed using a LEO 912 AB OMEGA electron microscope (Leo Elektronenmikroskopie, Oberkochen, Germany).

### Lipidome analyses

Lipidome analysis was performed using the Lipidyzer™ Platform from SCIEX. Plasma samples were spiked with Lipidyzer™ Internal Standards (SCIEX), following lipid extraction employing an adjusted 2-methoxy-2-methylpropane (MTBE) / methanol (MeOH) extraction protocol^10^. Lipids were concentrated and reconstituted in a 50 : 50 mixture of dichloromethane and MeOH containing 10 mM ammonium acetate. Differential mobility spectrometry and QTRAP^®^ system processing (QTRAP^®^ 5500; SCIEX) permitted separation and targeted profiling of lipid species. Lipids were quantified using Lipidyzer™ software (Lipidomics Workflow Manager; SCIEX) using specific multiple reaction monitoring transitions.

### Mass spectrometry

For homogenization, tissue was transferred into a 2 ml sample tube. Lysis buffer, 900 µl, HALT phosphatase inhibitor cocktail (Thermo Fisher), and a steel sphere were added and the tube contents were homogenized in a bead mill (TissueLyser II, Qiagen, Hilden, Germany) for 2 min at 25 Hz. The sample was incubated for 5 min at 95°C and 800 rpm and was ultrasonicated with 5 pulses at 30% of power. Cold MTBE, 750 µl, was added and the sample was shaken for 1 h at 4°C. To induce phase separation, 188 µl water with 0.1% ammonium acetate was applied. The extract was then centrifuged at 10,000 g for 5 min and the upper phase, containing lipids, was collected. To precipitate proteins, MeOH was added to the remaining lower phase in a 4:1 ratio, v/v MeOH/water. The mixture was incubated for 1 h at -20°C and then centrifuged for 12 min (13,000 g) at 4°C. The remaining pellet was suspended in lysis buffer. Bicinchoninic acid assays (Thermo Fischer) were performed for all samples following manufacturer instructions. For proteomics samples, the single-pot, solid-phase, sample-preparation protocol was followed with 20 µg as starting material as described above. For phosphoproteomics samples, 200 µg were used as starting material. After tryptic digestion, phosphopeptides were enriched using the High-Select Fe-NTA Magnetic Phosphopeptide Enrichment Kit (Thermo Fischer) according to manufacturer instructions. Chromatographic separation of peptides was achieved with a two-buffer system (buffer A, 0.1% formic acid [FA] in H_2_O; buffer B, 0.1% FA in acetonitrile [ACN]) on a nano – ultra-high-pressure liquid chromatograph (UHPLC; Dionex Ultimate 3000 UHPLC system, Thermo Fisher). Attached to the UHPLC was a peptide trap (100 µm x 20 mm, 100 Å pore size, 5 µm particle size, C18, Nano Viper, Thermo Fisher) for online desalting and purification, followed by a 25 cm C18 reversed-phase column (75 µm x 250 mm, 130 Å pore size, 1.7 µm particle size, peptide BEH C18, nanoEase, Waters). Peptides were separated using an 80 min method with linearly increasing ACN concentration from 2% to 30% over 60 min.

Tandem mass spectrometry (MS/MS) was conducted using a quadrupole-ion-trap-orbitrap mass spectrometer (Orbitrap Fusion, Thermo Fisher). Eluting peptides were ionized using a nano-electrospray ionization source with a spray voltage of 1,800 V and analyzed in data independent acquisition (DIA) mode. For each initial scan, ions were accumulated for a maximum of 240 milliseconds or until a charge density of 2 x 10^5^ ions (automatic gain control [AGC] target) was reached. Fourier-transformation based mass analysis of the data from the orbitrap mass analyzer was performed covering a mass range of m/z 400 – 1,200 with a resolution of 120,000 at m/z = 200. Fragmentation in DIA mode with m/z 12 isolation windows and m/z 1 window overlaps was performed within a mass range of m/z 400 – 800. Fragmentation was done with a normalized collision energy of 30% using higher energy collisional dissociation. An AGC target of 5 x 10^4^ ions or a maximum of 54 ms was set. Orbitrap resolution was set to 30 000 with a scan range from m/z 350-2000.

Liquid chromatography-MS/MS data were searched with the Sequest algorithm integrated into Proteome Discoverer software (v3.1, Thermo Fisher Scientific) against a reviewed mouse database, obtained in November 2023, that contained 17,289 entries, using Inferys 3.0 fragmentation as prediction model. Carbamidomethylation was set as a fixed modification for cysteine residues. The oxidation of methionine was allowed as a variable modification. For phosphoproteomics samples, phosphorylation of serine, threonine, and cysteine was set as a variable modification. A maximum number of one missing tryptic cleavage was set. Peptides between 7 and 30 amino acids were considered. A strict cutoff (false discovery rate (FDR) < 0.01) was set for peptide identification. Quantification was performed using the CHIMERYS algorithm (MSAID) based on fragment ions. Protein abundances were log2 transformed and normalized to the column median before statistical analysis.

### Western blot

Cultured cells were washed twice with cold phosphate buffered saline (PBS) and scraped off in 1 ml of PBS buffer. The mixture was centrifuged for 5 min at 650 xg and 4°C. The supernatant was removed and resuspended in 50 µl cell lysis buffer (50 mM Tris–HCl, pH 8.0; 150 mM NaCl; 1% Nonidet P-40; 0.5% Na-deoxycholate; 5 mM EDTA, 0.1% sodium dodecyl sulfate [SDS]) supplemented with complete Mini Protease Inhibitors and PhosStop (Roche). Organs were harvested and homogenized directly in supplemented lysis buffer. Clarification of cell lysates and organ lysates was performed by centrifugation for 10 min at 14,000 rpm and 4°C. Supernatants were supplemented with sample buffer and proteins were separated on SDS-polyacrylamide gels and transferred to polyvinylidene fluoride membranes using the Transblot Turbo Transfer System (Bio-Rad Laboratories). Following blocking (20 mM Tris–HCl, pH 7.4; 150 mM NaCl; 0.1% Tween-20; 5% non-fat dry milk) and washing (20 mM Tris–HCl, pH 7.4; 150 mM NaCl; 0.1% Tween-20), membranes were incubated in primary antibody solution (20 mM Tris–HCl, pH 7.4; 150 mM NaCl; 0.1% Tween-20; 5% BSA or 5% non-fat dry milk) overnight containing the appropriate antibodies. Membranes were washed and incubated with donkey anti-rabbit IgG horseradish peroxidase secondary antibody (GE Healthcare; no. NA934V; 1:7,500 dilution) or with sheep anti-mouse IgG horseradish peroxidase secondary antibody (GE Healthcare; no. NA931V; 1:7,500 dilution) for 1 h. After final washing, protein signals were visualized using the ChemiDoc MP Imaging System (Bio-Rad Laboratories).

### Exome sequencing and variant validation

Genomic DNA extracted from blood samples was subjected to trio exome sequencing. Coding DNA fragments were enriched with a SureSelect Human All Exon 50Mb V5 Kit (Agilent) and libraries were sequenced on a HiSeq2500 platform (Illumina). Reads were aligned to the human reference genome (UCSC GRCh38/hg19) using the Burrows-Wheeler aligner (v.0.5.87.5). Genetic variation was detected using SAMtools (v.0.1.18), PINDEL (v. 0.2.4t), and ExomeDepth (v.1.0.0). Impact of predicted amino acid substitutions on protein function was assessed by the pathogenicity prediction tools CADD, M-CAP, and ClinPred. Variants were validated by Sanger sequencing. Primer pairs were designed to amplify selected coding exons of the candidate gene. Amplicons were directly sequenced using an ABI BigDye Terminator Sequencing kit (Applied Biosystems) and a capillary sequencer (ABI 3500, Applied Biosystems). Sequence electropherograms were analyzed using Sequence Pilot software (JSI Medical Systems).

### Statistical methods

Data are displayed as mean ± standard error of mean comparisons of groups that were examined using Student’s *t*-test or the Mann-Whitney *U* test. Comparisons of 3 or more groups were analyzed using ANOVA. Perseus (Version 2.0.11.0), GraphPad Prism and Microsoft Excel were used for all statistical analyses.

## Data Availability

All data produced in the present study are available upon reasonable request to the authors.

## Acknowledgments

The authors thank Katrin Rading, Laura Ehlen, Verena Rickassel, and Meike Kröger for excellent technical support. C.S. was supported by grants from the Deutsche Forschungsgemeinschaft (DFG; SCHL2276/2-1, 450149205-TRR333/1). A.W. and J.H. were supported by grants from the Deutsche Forschungsgemeinschaft (450149205-TRR333/1). H.S. was supported by grants from the DFG (INST 337/15-1, INST 337/16-1, INST 152/837-1, INST 152/947-1 FUGG, and SCHL 406/21-1). We thank the Core Facility Mass Spectrometric Proteomics as part of the Technology Platform Mass Spectrometry at University of Hamburg and University Medical Center Hamburg-Eppendorf for support with mass spectrometric measurements and analysis funded by the DFG. B.W and D.K. are members of the European Reference Network for Rare Hereditary Metabolic Disorders (MetabERN) - Project ID No 739543.

---

**Figure S1.**
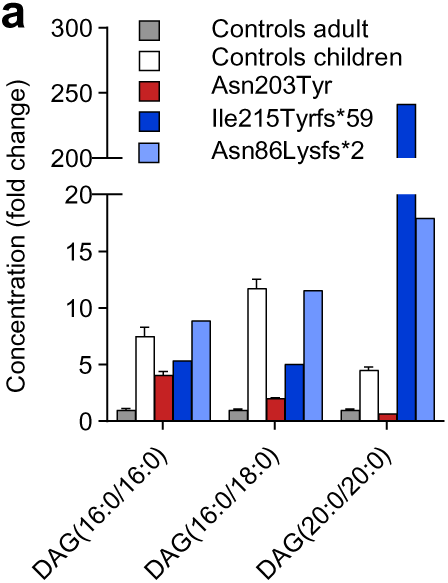
(a) Plasma concentration -fold changes of diacylglycerides (16:0/16:0), (16:0/18:0), and (20:0/20:0) in individuals with *TXNIP* variants yielding TXNIP p.Asn203Tyr, TXNIP p.Asn86Lysfs*2, and TXNIP p.Ile215Tyrfs*59 (n=1 each) versus adult (n=9) and children controls (n=3).

