## Supplemental information for "TXNIP is a critical regulator of human cardiometabolic health"

**Supplementary information**

| **Content** | **Page** |
| --- | --- |
| Medical history of individuals #1-6 | 1 |
| Supplemental references | 7 |

***Individual #1***

A man in his 40's presented with cardiomyopathy of unknown origin, exercise intolerance, and muscle weakness. Recurrent lactatemia and elevated serum creatine kinase (CK) activity levels were documented. Exercise intolerance had been present since youth. Cardiomyopathy was discovered in his 20's on routine check-up. Upper-extremity muscle weakness began in his 40's. Lactate values were 14.2 mmol/l at maximum and 5.6 mmol/l at minimum. He reported a single hypoglycemic event (36 mg/dl) that required medical attention.

He was born to healthy, non-consanguineous parents. One parent has coronary heart disease, arterial hypertension, rheumatism, and leukaemia. The medical history of the other parent is unremarkable. A younger sibling is healthy. An older sibling died from acute decompensation of cardiac insufficiency in his 30's only days after diagnosis of a previously unrecognized cardiomyopathy.

The individual's childhood development was unremarkable. Exercise intolerance became evident during physical education in school. Despite regular exercise, the individual had difficulties building endurance and muscle strength. He reported that earlier testing had shown slightly elevated serum CK-MB values. These were not investigated. His cardiomyopathy was of mixed hypertrophic and dilatative form. During follow up, ejection fraction (EF) in his 40's was 31% at minimum, leading to initiation of treatment for cardiac insufficiency. In his 40's, permanent chest pain developed, accompanied by muscle pain and weakness of upper-extremity muscles. At the time of presentation, echocardiography showed improved EF to 41% upon heart failure therapy as well as moderate left ventricular hypertrophy. He also had left bundle branch block and arterial hypertension. On tertiary-care referral, values for CK-MB (404 U/l) and lactate (10.2 mmol/l) were elevated. During ergometry, lactate values rose to 14.2 mmol/l. Cardiac stress magnetic resonance imaging found no signs of myocardial stress-induced ischemia. Histologic examination of endomyocardial-biopsy material found hypertrophied myocytes and mild interstitial fibrosis, but no signs of hypertrophic or dilatative cardiomyopathy. Hyperlactatemia prompted ultrastructural study for evidence of mitochondriopathy. Mitochondria were abnormal in configuration and size, with slightly irregular cristae.

Exome sequencing revealed a 334 kb TAR (thrombocytopenia – absent-radius syndrome) deletion in 1q21.1, spanning *TXNIP*, on one allele and a *TXNIP* c.253G>T, p.Glu85* nonsense variant on the other allele, indicating biallelic TXNIP deficiency.

Besides cardiac and muscular issues, the individual suffers from memory difficulties. Since his 40's, he has sometimes forgot appointments and the names of long-known colleagues and friends. He reports gender dysphoria and has been diagnosed as infertile owing to abnormal spermatogenesis.

The individual self-initiated a sugar-free diet for several weeks per year with self-reported increase in well-being, muscular strength, and mobility.

***Individual #2***

Individual #2 died in his 30's in multi-organ failure following acute heart failure caused by a previously undetected cardiomyopathy. Genetic testing revealed a 334 kb TAR deletion in 1q21.1, spanning *TXNIP*, on one allele and a *TXNIP* c.253G>T, p.Glu85* nonsense variant on the other allele.

He described himself as healthy before the onset of acute illness, with self-referral to an emergency room due to chest pain. Echocardiography revealed a substantially reduced biventricular EF (10%) and a thrombus in the dilated left ventricle. Initial blood-gas analysis found a lactate level of 15 mmol/l. On the same day he was admitted to a cardiac-surgery intensive care unit (ICU) where a left ventricular assist device was placed and veno-arterial extracorporeal membrane oxygenation (ECMO) for right ventricular assistance was begun. Acute renal failure was treated with veno-venous hemodialysis. Lactate values rose to > 31 mmol/l leading to severe metabolic acidosis, that was only partial adjustable by tris-buffer administration. In the course of ICU treatment, a CT scan revealed cerebrovascular hemorrhage, with elevated intracranial pressure. This was treated by hemicraniectomy. Subsequently, he had multi-organ failure with persistent lactatemia and global hypoperfusion of the brain despite use of cardiac support systems and maximal intensive-care therapies. Only a few days after initial presentation in the emergency room, he was in end-stage multi-organ failure. Death supervened when supportive care was withdrawn. A post-mortem report of molecular-pathologic findings in a heart biopsy specimen taken during support-device placement surgery detected parvovirus B19 DNA, indicating that concurrent myocarditis might have aggravated his underlying severe cardiomyopathy.

***Individual #3***

This boy is the 4^th^ child of consanguineous parents. His 3 siblings and his parents were well. During pregnancy, corpus callosum agenesis and signs suggesting hydrops fetalis were noted. He was born at 38+2 weeks by cesarean section due to breech position. Generalized hypotonia, hepatomegaly, and facial dysmorphism were noted at birth, as were a cataract of the left eye and a patent foramen ovale without hemodynamic consequences. A congenital lactic acidosis (max 9 mmol/l, ref < 2 mmol/l) decreased after the neonatal period to 3-4 mmol/l. Two attempts at 3:1 ketogenic dietary therapy were stopped due to hyperlipidemia. At lst follow- up, he exhibits generalized hypotonia and severe delay in all areas of development. He can walk ~300 meters with assistance and communicates orally with sounds and a handful of meaningful words. He is a friendly and interactive child with no behavioural issues.

***Individual #4***

This woman first presented in her 20's with an episode of abdominal pain and emesis after a fasting period. Ketotic and lactic acidosis accompanied hypoglycemia. Fasting serum triglyceride values were elevated (325 mg/dl, ref. <150 mg/dl), as were serum levels of creatinine (1.87 mg/dl) and uric acid (13.4 mg/dl). CK values were not elevated. Echocardiography showed no signs of hypertrophic cardiomyopathy or impaired ventricular function. Symptoms as well as lactic acidosis and ketosis resolved with parenteral glucose infusion. Serum triglycerides remained slightly elevated (189 mg/dl, ref. <150 mg/dl), but kidney function normalized. Several months earlier, she had been admitted to hospital elsewhere with similar symptoms and was similarly treated. She had been diagnosed with essential arterial hypertension in her teens.

She suffered from multiple calcium oxalate kidney stones. Previous investigation had identified chronically elevated CK values (400-500 IU/L) and asymptomatic low glucose levels (~ 45 mg/dl). Serum transaminase activities were within normal ranges and serum lipid values fluctuated between normal and moderately elevated levels. The individual’s 2 siblings and one of their parents are healthy. No information is available on her other parent.

Childhood development was unremarkable. She repeatedly complained of muscle weakness, but neurologic examination identified no abnormality. An electromyographic study was not performed. Self-reported hearing impairment was not confirmed by an otolaryngologist. Exome sequencing revealed a biallelic frameshift variant (c.[257dupA]; p.Asn86Lysfs*2) in *TXNIP*.

Use of a subcutaneous glucose sensor revealed prolonged low glucose levels, especially during night-time. The individual was advised to take 40 g of starch before going to bed as well as to avoid longer fasting periods. Two additional episodes of ketotic and lactic acidosis have occurred after fasting or during illness. Her status is being monitored annually. Slight elevations persist in levels of lactate, serum lipids (mostly LDL cholesterol), and CK.

***Individual #5***

A man in his 20's presented with progressive exercise-induced muscle weakness, arterial hypertension, and hypertrophic cardiomyopathy. His muscular symptoms began in early adulthood, while his cardiovascular issues had been present since late childhood. He was the second of 4 children born to healthy consanguineous parents

One parent died in his 60's of cardiomyopathy of undetermined etiology, while the other parent is in good health. His 3 siblings and his children are healthy.

His childhood development was unremarkable. The first symptom reported was tachycardia in his teens; elevated blood pressure was identified, with echocardiography revealing a bicuspid aortic valve and mild aortic-valve insufficiency. In his teens, hypertrophic cardiomyopathy and arterial hypertension were diagnosed. Antihypertensive medication was administered. A few years later, he noted a gradual decline in muscle strength, with progressively increasing weakness in hands and legs for approximately one year before initial presentation in his 20's. He did not then report cramps or impairment of sensation; cramps in his hands and legs developed later, with tingling paraesthesia. He described a sensation of dizziness and fainting on several occasions. Repeated blood glucose measurements gave results were within the low-normal or hypoglycemic range. A clinical fasting test showed asymptomatic hypoglycemia (glucose 52 mg/dl). Two seizures occurred, both in the context of hunger or stress. CK and lactate values were chronically high (respectively 500-1200 IU/L and 3.5-5 mmol/L), with lactic acidosis. Transaminase-activity and plasma lipid values fluctuated between normal and moderately elevated.

A muscle biopsy revealed excessive accumulation of both glycogen and abnormal lipid deposits. A liver biopsy showed moderate accumulation of both lipids and glycogen, as well as irregular mitochondrial structure. Exome sequencing revealed the biallelic variant c.607A>T, p.Asn203Tyr, in *TXNIP*.

Since his 20’s, the individual had several episodes of gout. He therefore began treatment with allopurinol. In his 20's, a surgical procedure was performed to remove a knee cyst.

in his 30's, he developed critical ischemia of both legs, owing to occlusion of the popliteal arteries by thrombus embolized from within the left ventricle. He underwent intraluminal lysis and embolectomy. Impaired wound healing was successfully treated with vacuum-assisted closure. Warfarin was begun to provide anticoagulation. During this hospitalization, lactate and CK levels were high (respectively 3.6-10.5 mmol/L and 352-651 IU/L).

Cardiac function remained stable for years, with slight to moderate impairment observed. In his 30's, he presented with palpitations, chest pain, and dyspnea. An absolute tachyarrhythmia (TAA) accompanied markedly reduced cardiac function, characterized by an EF of 10–15%, with acute decompensation manifest as low-output failure. Administration of the positive inotropic agent levosimendan permitted cardiac recompensation and stabilized cardiac function. The TAA was treated by electrocardioversion, followed by intensification of his pharmacologic heart-insufficiency regimen. During this hospitalization serum lactate levels ranged between 3.4 and 7.6 mmol/L. One month later, TAA recurred. It was again treated by cardioversion. Amiodarone was begun, but was withdrawn due to QTc-time prolongation. Pulmonary-vein isolation was performed to prevent further TAA. As cardiac function continued to deteriorate (EF 10%) and lactate levels ranged between 6.0 and 11.7 mmol/L, recurrence of low-output heart failure was suspected. Levosimendan administration did not lower lactate levels. He complained of nausea and discomfort; elevated transaminase activities and spontaneous dyscoagulation prompted suspicion of liver-function impairment. Liver sonography did not reveal any evidence of parenchymal damage. A slight increase in kidney retention parameters indicated impaired renal function. Metabolic decompensation was hypothesized. Stabilization was observed upon administration of fluids, parenteral alimentation, and coenzyme Q10, with improvements in organ function parameters as well as in coagulation. Concentrations of lactate also fell (4.1 mmol/l). During this hospitalization, CK levels were elevated (571 IU/L). Cardiac function recompensated but did not improve with medical treatment, so implantation of a cardiac resynchronization therapy (CRT) defibrillator was scheduled to prevent sudden cardiac death and to improve cardiac function. Evaluation for heart transplantation was offered but was declined on religious grounds. During CRT-device implantation, he once again developed arrhythmia, with a cardiac arrest shortly after the intravenous induction of anaesthesia (fentanyl / propofol). Resuscitative measures included implantation of a mechanical cardiac support pump and initiation of ECMO. Progressive cardiogenic shock nonetheless developed, with death from multi-organ failure several days later.

***Individual #6***

This boy is the first child of healthy consanguineous parents. Parts of his medical history have been described recently.^1^ Pregnancy was complicated by polyhydramnios. Due to breech position, cesarean section was performed at 38+6 weeks gestational age. He was transferred from the birth hospital to a tertiary-care hospital because of floppiness, recurrent hypoglycemia, and lactic acidosis. In his first postnatal days, the lowest measured concentration of glucose was 30 mg/dl, with lactate values around 6.5 mmol/l and pH 7.11. A mitochondrial disorder was suspected. The individual developed hypertriglyceridemia (up to 364 mg/dl). He was stable in the first days after admission in transfer. pCO2 levels rose as high as 70 mmHg (due to bradypnea) and metabolic acidosis developed. Glucose was given by vein (10 mg/kg/min) to stabilize blood glucose concentrations. Lactate concentrations also stabilized but remained elevated at ~ 3-4 mmol/l. Muscular hypotonia did not resolve. Liver and muscle biopsies were conducted, as was a skin punch biopsy for cultivation of fibroblasts. After some days, glucose and lactate concentrations were stable on regular feeds. He was transferred to a peripheral hospital.

Exome sequencing of DNA from peripheral blood leukocytes revealed homozygosity for a nucleotide insertion in *TXNIP* (c.642_642insT, p.I215Yfs*59) causing a missense mutation, followed by a frameshift with a premature stop codon after 59 amino acids and yielding a truncated TXNIP variant. Each parent was heterozygous for the same mutation.

Psychomotor development was delayed, with increasingly frequent epileptic seizures beginning in the first years of life (up to 3-4 per month, controlled with levetiracetam, topiramate, and lamotrigine). In response to stress, such as infections, the individual suffered from recurrent episodes of hypoglycemia. Currently he is metabolically stable, with marked developmental delay that has autistic features.

**Individuals #7, #8, #9**: Adapted from Katsu-Jimenez et al.^2^
